# Engagement in leisure activities and depression in older adults in the United States: Longitudinal evidence from the Health and Retirement Study

**DOI:** 10.1101/2021.04.15.21255495

**Authors:** Jessica K Bone, Feifei Bu, Meg Fluharty, Elise Paul, Jill K Sonke, Daisy Fancourt

**Author notes:** Corresponding author UCL Research Department of Behavioural Science and Health, 1-19 Torrington Place, London WC1E 7HB. Author contributions JKB, FB, and DF designed the study. JKB and FB conducted the analysis and JKB drafted the manuscript. JKB, FB, MF, EP, JKS, and DF contributed to the writing, made critical revisions, and approved the final manuscript.

## Abstract

**Objectives:** Receptive cultural engagement (e.g., attending theaters and museums) can reduce depression in older adults. However, whether specific participatory leisure activities are associated with lower rates of depression remains unknown. We aimed to test whether engagement in a diverse range of leisure activities, all of which could involve artistic or creative elements, was associated with concurrent and subsequent depression.

**Methods:** Using longitudinal data from 19,134 participants aged over 50 in the Health and Retirement Study, engagement in leisure activities was measured every four years, and depression every two years, between 2008 and 2016. Leisure activities included: reading books, magazines, or newspapers; writing; baking/cooking something special; making clothes, knitting, or embroidery (sewing); working on hobbies/projects; going to sport, social, or other clubs; and attending non-religious organization meetings. A score of three or more on the Center for Epidemiologic Studies Depression Scale indicated depression. We fitted population-averaged panel data models using generalized estimating equations with a logit link.

**Results:** Engaging in some leisure activities, such as clubs, hobbies/projects, and baking/cooking was associated with reduced depression, independent of confounders. Concurrently, spending time on hobbies/projects (monthly OR = 0.80, 95% CI = 0.72–0.88; weekly OR = 0.81, 95% CI = 0.73–0.89) and clubs (monthly OR = 0.85, 95% CI = 0.77–0.94; weekly OR = 0.78, 95% CI = 0.69–0.88) was associated with lower odds of depression versus not engaging. Longitudinally, the odds of depression two years later were reduced amongst people engaging in weekly baking/cooking (OR = 0.85, 95% CI = 0.75– 0.95), hobbies (OR = 0.81, 95% CI = 0.71–0.92), and clubs (OR = 0.82, 95% CI = 0.71–0.94). Writing, reading, sewing, and attending non-religious organizations were not consistently associated with depression.

**Discussion:** Engagement in some leisure activities is associated with reduced odds of depression. We should consider how older adults can be supported to actively participate in leisure activities as health-promoting behaviors.

## Introduction

In the United States (US), there is currently unprecedented growth in the number of older adults. By 2060, adults aged 65 and above are projected to form 23% of the American population (Vespa et al., 2020). Among this aging population, depression is one of the most common mental health problems, with point prevalence estimates ranging from 11% to 15% (Steffens et al., 2009; Zivin et al., 2010). Despite this, depression is commonly undiagnosed in older adults and is often undertreated (Rodda et al., 2011). As well as causing functional impairments in daily life, depression is associated with increased risks of cardiovascular disease, dementia, diabetes, stroke, and both specific and all-cause mortality (Cuijpers and Smit, 2002; Knol et al., 2006; Leonard, 2017; Pan et al., 2011; Van der Kooy et al., 2007). Additionally, identifying ways to reduce or prevent depression is particularly important in older age, when close social ties may be lost, people are more likely living alone, many have low income, and progressive age-related health problems may limit people’s activities (World Health Organization, 2015).

Leading a socially engaged life is thought to be important for successful ageing, which includes maintaining close relationships as well as being involved in activities that are meaningful and provide purpose, such as leisure activities (Rowe and Kahn, 1997). Leisure activities are voluntary non-work activities that are engaged in for enjoyment (Hills and Argyle, 1998), ranging from hobbies to participating in the arts to taking part in sports or exercise groups. There is extensive evidence that participating in leisure activities can support both the prevention and management of depression in older adults (Adams et al., 2011; Dupuis and Smale, 1995; Fancourt et al., 2020a; Fancourt and Tymoszuk, 2019; Glass et al., 2006; Hong et al., 2009; Katz and Yelin, 2001; Lampinen et al., 2006; Nimrod et al., 2012). This may be because leisure activities involve a range of active ingredients that are health promoting, including providing opportunities for creative expression, aesthetic pleasure, cognitive stimulation, sensory activation, evocation of emotion, and social interaction (Dunphy et al., 2019; Fancourt and Finn, 2019). These ingredients then activate a range of mechanisms of action that may reduce depression, including psychological processes (e.g. improving self-esteem and mood), biological processes (e.g. reducing levels of stress hormones and inflammation), social processes (e.g. reducing loneliness and improving communication), and behavioral processes (e.g. enhancing agency and motivation to engage in other health behaviors; Fancourt et al., 2021).

Cultural, arts, and creative activities represent one domain of leisure activity that may be particularly beneficial for reducing depression in older adults (Adams et al., 2011; Dupuis and Alzheimer, 2008). These cultural activities are commonly split into those that are receptive, involving art that has been created and is now experienced by an audience, and those that are participatory, requiring the creation of and participation in the arts (Fancourt et al., 2021; Fancourt and Finn, 2019; Tymoszuk et al., 2021). Although there is strong evidence from large observational studies that receptive cultural engagement (such as going to the theatre and museums) is associated with lower rates of depression in older adults (Elsden and Roe, 2020; Fancourt and Steptoe, 2019; Fancourt and Tymoszuk, 2019; Ryu and Heo, 2018), these activities may not be accessible to all. For older adults living in the community, receptive cultural engagement may be subject to more socioeconomic barriers than other leisure activities (Bone et al., 2021), and inequalities in access may increase with age (Fluharty et al., 2021). More than one in three older adults in the US have reported some difficulty attending cultural venues (Rajan and Rajan, 2017), which may be a result of cultural equity and relevance (Bone et al., 2021).

In contrast, a wide range of participatory creative and arts-based leisure activities can be done in the home, many of which are inexpensive or free. Despite these activities being more accessible than receptive cultural activities, there is less evidence on the associations between them and depression in older adults. Previous research has typically grouped a range of participatory creative activities into total engagement (Glass et al., 2006; Katz and Yelin, 2001; Lampinen et al., 2006), an approach that has been criticized for combining conceptually distinct pursuits (Kerby and Ragan, 2002; Ritchey et al., 2001; Ryu and Heo, 2018). Identifying activities that are, and are not, associated with depression will allow us to understand the characteristics of activities providing the most benefits. For example, we know that the social element of activities is usually important and protective (Adams et al., 2011; Dadswell et al., 2017; Dupuis and Alzheimer, 2008; Ryu and Heo, 2018), but we do not know whether creative or artistic activities provide additional benefits over and above social engagement. When examining more specific forms of leisure, one longitudinal study found that actively engaging in creative activities, but not cultural attendance, was associated with better mental health in a large sample of adults aged 16 to 101 years (Wang et al., 2020). Engagement in hobbies and crafts is also associated with reductions in subsequent depression in older adults (Dupuis and Smale, 1995; Fancourt et al., 2020a). However, these studies included predominantly White British samples, making generalizability across different nationalities and races/ethnicities difficult. Furthermore, only one indicator of engagement was used, meaning different activities could not be compared.

In this study, we used a large nationally representative cohort of older adults in the US (the Health and Retirement Study; Sonnega et al., 2014) to explore associations between engagement in a range of leisure activities and concurrent and subsequent depression. We measured seven activities that are commonly done by older adults: reading books, magazines or newspapers; writing (e.g., letters, stories, or journal entries); baking or cooking something special; making clothes, knitting, or embroidery (sewing); working on a hobby or project; going to a sport, social, or other club; and attending meetings of non-religious organizations (political, community, or other interest groups). Although not traditionally labelled as ‘arts’, we included this diverse range of activities as they could all include artistic or creative engagement, potentially sharing similar properties of creative skill and imagination. These activities combine complex and distinct elements, differing in specificity as well as the extent to which they could involve creativity and whether they are likely to entail social interactions. We therefore aimed to investigate whether these activities were differentially associated with depression, furthering our understanding of the characteristics of leisure activities that are important for preventing or reducing depression in older adults. We hypothesized that more frequent engagement in creative activities would be associated with lower odds of concurrent and subsequent depression.

## Methods

### Sample

Participants were drawn from the Health and Retirement Study (HRS); a nationally representative study of more than 37,000 individuals over the age of 50 in the US (Sonnega et al., 2014). The initial cohort was first interviewed in 1992 and followed-up every two years, and the sample is replenished with younger cohorts every six years (Sonnega et al., 2014). We used several RAND HRS public datasets (Longitudinal File 2016 (V2); Detailed Imputations File 2016 (V2); 2008-2016 Fat Files).

We used data from HRS waves at which engagement in leisure activities was consistently measured (2008-2016). At each wave, a rotating random 50% subsample of participants were invited to an enhanced interview and given a Leave-Behind Psychosocial and Lifestyle Questionnaire (LBQ) to complete and return by mail, which included questions on leisure activities (Smith et al., 2017). Participants were eligible to complete the LBQ in 2008, 2012, and 2016 or in 2010 and 2014. Response rates in each year varied from 62% to 85%. In 2008, 8,296 participants were invited to complete this questionnaire, and 7,073 (85%) returned it. In 2010, 11,213 were eligible and 8,332 (74%) participated. In 2012, 10,079 were eligible and 7,412 (74%) participated. In 2014, 9,549 were eligible and 7,541 (79%) participated. In 2016, 10,238 were eligible and 6370 (62%) participated. In total, 20,643 individuals completed between 1 and 4 waves (mean=1.8 waves). Restricting this sample to participants with complete data on depression and the LBQ in at least one wave produced a final sample of 19,134 participants and 33,880 observations (1.8 repeated measures per person, range 1-3). See Figure S1 for a flowchart illustrating the sample selection process.

All participants gave informed consent, and this study has approval from the University of Florida (IRB201901792) and University College London Research Ethics Committee (project 18839/001).

### Outcome

Depression was measured at every wave with the modified eight-item Center for Epidemiologic Studies Depression Scale (CES-D; Steffick, 2000; Turvey et al., 1999). This self-report measure can identify people at risk of developing depression and has comparable psychometric properties to the twenty-item CES-D (Turvey et al., 1999). Total possible score ranges from 0 to 8, with higher scores indicating more severe symptoms. The CES-D had good internal consistency across waves (Cronbach’s alpha ranged from 0.81 to 0.82). We used a cut-off of three or more to indicate the presence of depression (Steffick, 2000; Turvey et al., 1999).

### Exposures

The LBQ included a 20-item Social Participation - Social Engagement scale, which included questions on a wide range of leisure activities (Smith et al., 2017). We selected seven items from this questionnaire that were measured consistently from 2008 to 2016 and could include elements of artistic and creative skill and imagination. Participants were asked how often they: 1) read books, magazines, or newspapers; 2) do writing (such as letters, stories, or journal entries); 3) bake or cook something special; 4) make clothes, knit, embroider, etc. (sewing); 5) work on a hobby or project; 6) go to a sport, social, or other club; and 7) attend meetings of non-religious organizations (political, community, or other interest groups). We collapsed responses into three categories, representing no engagement (never/not in the last month), monthly engagement (once a month/several times a month), and weekly engagement (once a week/several times a week/daily). Participants received these questions once every two waves as part of the rotating random 50% subsample invited to complete the LBQ (2008/2012/2016 or 2010/2014).

### Confounders

We included a range of demographic, socioeconomic, and health-related confounders. Demographic confounders were age (50-59, 60-69, 70-79, ≥89 years), gender (men, women), marital status (married [including cohabiting], unmarried), and race/ethnicity (White [including Caucasian], Black [including African American], Other [including American Indian, Alaskan Native, Asian or Pacific Islander, Hispanic, Other]). In the public HRS data, this variable indicated the race/ethnicity as which participants primarily identified and detailed information was removed to protect participant confidentiality.

Socioeconomic confounders were educational attainment (none, high school, college, postgraduate), employment status (employed, not working [including unemployed, temporarily laid off, disabled, homemakers], retired), total household income in quartiles (<$19,000, $19,000-$39,999, $40,000-$79,999, ≥$80,000), neighborhood safety (excellent/good, fair/poor), and social network composition (less diverse, more diverse). Social network composition was measured by asking participants if they had a spouse/partner, children, other immediate family, or friends. Those who did not have all of these social contacts were classified as part of a less diverse social network (Smith et al., 2017).

Health-related confounders included difficulties relating to activities of daily living (ADLs; none, one or more). Participants were asked whether, because of a health or memory problem, they had any difficulty with dressing, bathing or showering, eating, getting in or out of bed, and walking across a room (Bugliari et al., 2020). We also included difficulties relating to instrumental activities of daily living (IADLs; none, one or more), which were making phone calls, managing money, taking medications, shopping for groceries, and preparing hot meals. A variable describing chronic health conditions (none, one or more) indicated whether participants reported having diabetes, lung disease, cancer, heart conditions, high blood pressure, arthritis, complications from stroke, or other medical conditions. Finally, we included a measure of cognition in quartiles (0-7, 8-10, 11-12, 13-20), which was a summary of performance on immediate and delayed recall tests, with higher quartiles indicating better cognition (Bugliari et al., 2020).

Three confounders (gender, ethnicity, and education) were time-invariant, so a consistent value was assigned to participants across all waves. Time-varying confounders were included for each wave in which participants completed the LBQ.

### Statistical analyses

We first explored the sociodemographic characteristics of the sample at baseline. We then investigated whether engagement in leisure activities was associated with concurrent and subsequent depression. Concurrent models included activity engagement and depression measured simultaneously, with estimates averaged across all waves (2008-2016). Longitudinal models tested the association between activity engagement in one wave (2008-2014) with depression in the subsequent wave (2010-2016). We adjusted for depression in the previous wave (2008-2014), as depression at follow-up is related to both activity engagement and previous depression. This longitudinal model thus estimated the associations between leisure activity engagement and change in depression two years later.

We fitted population-averaged panel data models by using generalized estimating equations (GEE). This allowed us to include repeated measures, with waves clustered within individual. We used an exchangeable correlation matrix to optimize model power and efficiency, although GEE is relatively robust to the choice of correlation structure (Twisk, 2003). We modelled depression with a binomial distribution, logit link, and robust standard errors. All models are presented before and after adjustment for confounders.

We imputed missing data on exposures and confounders using multiple imputation by chained equations (White et al., 2011). We used logistic, ordinal, and multinomial regression according to variable type, generating 20 imputed data sets using all variables included in analyses. The results of imputed analyses did not vary to complete cases (Tables S2-S4), so findings from imputed models are reported. All analyses were weighted to account for non-response, age, gender, and race/ethnicity, making the sample representative of all non-institutionalized individuals in the US population aged over 50 (Ofstedal et al., 2011; Smith et al., 2017). Analyses were performed using Stata 16 (StataCorp, 2019).

#### Sensitivity analyses

We investigated whether the associations between activity engagement and depression differed according to gender or age group. We first included an interaction term between each leisure activity and gender in the concurrent and longitudinal models. Where there was evidence for an interaction, we examined the association separately for men and women. After collapsing age intro three categories (50-59, 60-69, ≥70 years), we repeated this approach for age group.

We used a cut-off of three or more on the CES-D to indicate depression, but a cut-off of four or more has also been used (Steffick, 2000). HRS included the Short Form Composite International Diagnostic Interview (CIDI-SF), which can be used to determine a probable diagnosis of a major depressive episode according to DSM criteria (Nelson et al., 1998; Steffick, 2000). We thus repeated analyses using the CES-D alternative threshold and the CIDI-SF. Finally, we repeated analyses with the addition of preliminary data from the 2018 HRS.

## Results

In total, 19,134 participants provided data on leisure activities and depression in at least one wave of HRS. After weighting, 55% of participants were women, 85% were White/Caucasian, and 47% were retired. As white people represent less than 80% of the US population (Gibson and Jung, 2002), this is an underrepresentation of other groups. Overall, 20% of the sample met diagnostic criteria for depression. The prevalence of depression differed according to several socioeconomic and health-related factors (Table 1), as did the frequency of engagement in leisure activities (Table S1).

**Table 1.**
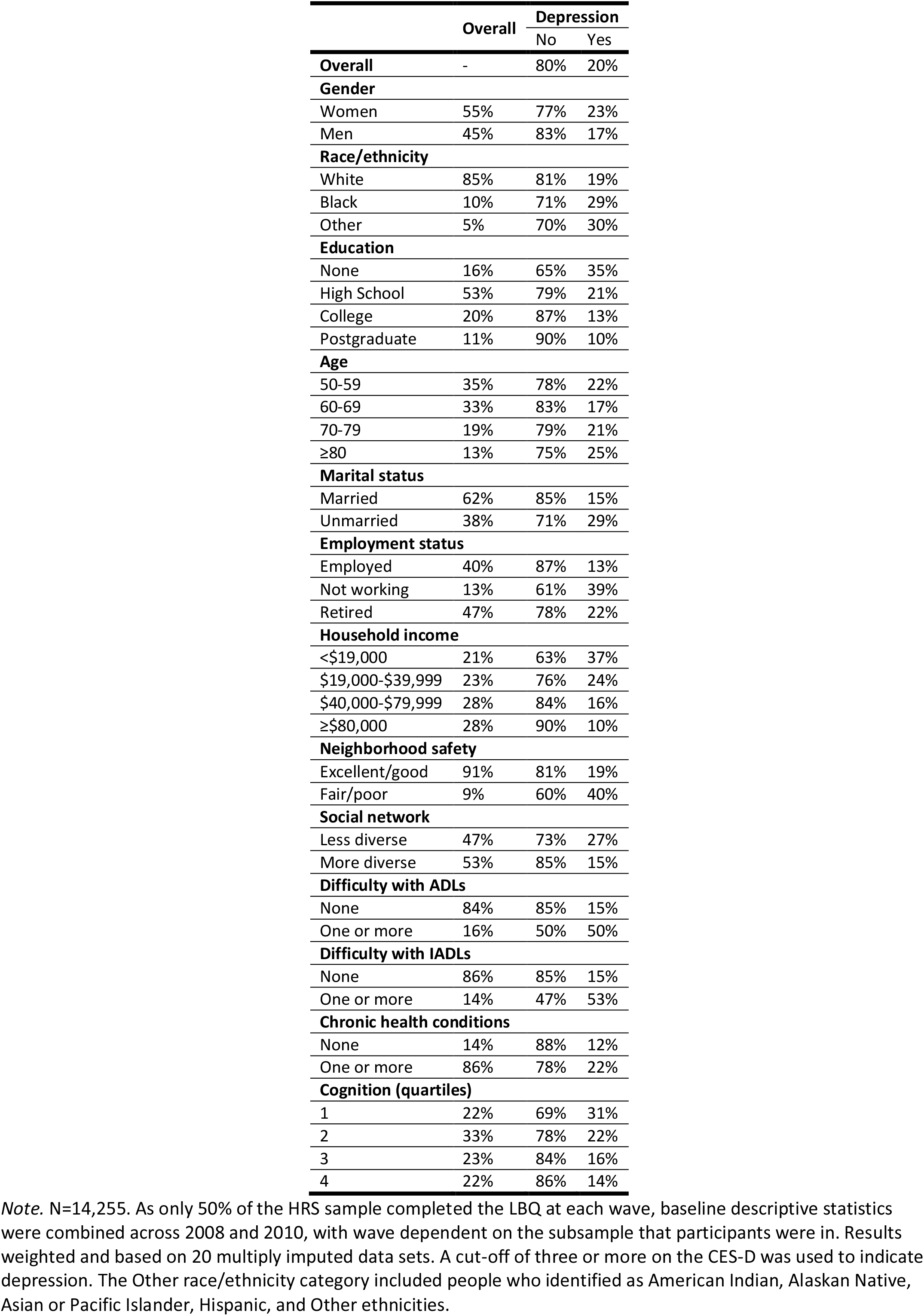
Demographic characteristics of the sample at baseline and percentage of the sample with depression at baseline according to demographic characteristics.

Frequency of engagement differed considerably across activities; 85% of participants read weekly, whereas only 4% attended meetings of non-religious organizations weekly (political, community, or other interest groups; Table 2). After reading (books, magazines, or newspapers), the most common activities were baking or cooking something special, working on a hobby or project, and writing (e.g., letters, stories, or journal entries).

**Table 2.** Frequency of leisure activities at baseline.

|  | Reading | Writing | Baking/cooking | Sewing | Hobby/project | Club | Organization |
| --- | --- | --- | --- | --- | --- | --- | --- |
| None | 6% | 61% | 31% | 86% | 43% | 64% | 81% |
| Monthly | 9% | 20% | 28% | 7% | 25% | 22% | 15% |
| Weekly | 85% | 19% | 41% | 7% | 32% | 14% | 4% |

*Note.* N=14,255. Results weighted and based on 20 multiply imputed data sets. Leisure activities were defined as: reading books, magazines, or newspapers; writing (e.g., letters, stories, or journal entries); baking or cooking something special; making clothes, knitting, or embroidery (sewing); working on a hobby or project; going to a sport, social, or other club; and attending meetings of non-religious organizations (political, community, or other interest groups).

### Concurrent associations

Before adjusting for confounders, we found evidence that more frequent reading, baking/cooking something special, working on hobbies/projects, and attending sport social or other clubs were associated with lower odds of depression (Table 3). In contrast, more frequent sewing (making clothes, knitting, or embroidery) and non-religious organizations (political, community, or other interest groups) were associated with increased odds of depression. After adjustment for confounders, there was only evidence that engaging in hobbies/projects and clubs were associated with depression. Compared to participants who did not spend time on a hobby/project, those doing hobbies/projects monthly had 20% (95% CI=0.72-0.88) lower odds of depression. This association was very similar for those who reported doing hobbies/projects weekly (OR=0.81, 95% CI=0.73-0.89). Engagement in clubs showed more of a dose-response relationship with depression. Monthly attendance was associated with a 15% (95% CI=0.77-0.94) reduction in odds of depression and weekly attendance was associated with a 22% (95% CI=0.69-0.88) reduction compared to not attending clubs.

**Table 3.** Concurrent models testing associations between frequency of leisure activities and the odds of depression.

|  | Model 1: Unadjusted |  |  | Model 2: Adjusted |  |  |
| --- | --- | --- | --- | --- | --- | --- |
|  | OR | 95% CI | p value | OR | 95% CI | p value |
| **Reading** |  |  |  |  |  |  |
| Monthly | 0.72 | 0.62-0.83 | <0.001 | 1.00 | 0.85-1.17 | 0.959 |
| Weekly | 0.55 | 0.49-0.62 | <0.001 | 0.92 | 0.80-1.05 | 0.206 |
| **Writing** |  |  |  |  |  |  |
| Monthly | 1.07 | 0.98-1.17 | 0.123 | 1.03 | 0.93-1.14 | 0.531 |
| Weekly | 0.98 | 0.89-1.08 | 0.714 | 0.98 | 0.88-1.09 | 0.725 |
| **Baking/cooking** |  |  |  |  |  |  |
| Monthly | 0.93 | 0.85-1.01 | 0.103 | 0.98 | 0.89-1.08 | 0.689 |
| Weekly | 0.84 | 0.77-0.92 | <0.001 | 0.91 | 0.82-1.01 | 0.084 |
| **Sewing** |  |  |  |  |  |  |
| Monthly | 1.09 | 0.94-1.26 | 0.242 | 0.92 | 0.79-1.08 | 0.314 |
| Weekly | 1.34 | 1.16-1.54 | <0.001 | 1.05 | 0.89-1.23 | 0.586 |
| **Hobby/project** |  |  |  |  |  |  |
| Monthly | 0.62 | 0.57-0.68 | <0.001 | 0.80 | 0.72-0.88 | <0.001 |
| Weekly | 0.62 | 0.57-0.68 | <0.001 | 0.81 | 0.73-0.89 | <0.001 |
| **Club** |  |  |  |  |  |  |
| Monthly | 0.68 | 0.62-0.74 | <0.001 | 0.85 | 0.77-0.94 | 0.001 |
| Weekly | 0.61 | 0.54-0.68 | <0.001 | 0.78 | 0.69-0.88 | <0.001 |
| **Organization** |  |  |  |  |  |  |
| Monthly | 0.91 | 0.81-1.01 | 0.086 | 0.98 | 0.87-1.10 | 0.716 |
| Weekly | 1.21 | 1.01-1.44 | 0.034 | 1.20 | 0.98-1.46 | 0.073 |

*Note.* N=19,276. For all activities, no engagement was the reference category. Model 2 was adjusted for gender, race/ethnicity, education, age, marital status, employment status, household income, neighborhood safety, social network, difficulty with ADLs and IADLs, chronic health conditions, and cognition. Results weighted and based on 20 multiply imputed data sets. Bold text indicates $p < 0.05$ .

### Longitudinal associations

Before adjusting for confounders, we found evidence that more frequent reading, baking/cooking something special, hobbies/projects, sport social or other clubs, and non-religious organizations (political, community, or other interest groups) were associated with lower odds of depression two years later (Table 4). As in the unadjusted concurrent model, more frequent sewing (making clothes, knitting, or embroidery) was associated with higher odds of subsequent depression in the unadjusted model. However, in the fully adjusted model, there was only evidence that baking/cooking special, hobbies/projects, clubs, and non-religious organizations were associated with subsequent depression. Participants who baked/cooked weekly had 15% (95% CI=0.75-0.95) lower odds of depression two years later and participants who did hobbies/projects weekly had 19% (95% CI=0.71-0.92) lower odds of subsequent depression. Both weekly and monthly club attendance were associated with a 18% (monthly 95% CI=0.73-0.93; weekly 95% CI=0.71-0.94) reduction in the odds of depression two years later. Finally, there was weak evidence that monthly (but not weekly) non-religious organization attendance was associated with lower odds of subsequent depression (OR=0.86, 95% CI=0.75-0.99).

**Table 4.** Longitudinal models testing associations between frequency of leisure activities and the odds of depression in the subsequent wave (two years later).

|  | **Model 1: Unadjusted** |  |  | **Model 2: Adjusted** |  |  |
| --- | --- | --- | --- | --- | --- | --- |
|  | OR | 95% CI | p value | OR | 95% CI | p value |
| **Reading** |  |  |  |  |  |  |
| Monthly | 0.72 | 0.61-0.86 | <0.001 | 1.03 | 0.83-1.26 | 0.811 |
| Weekly | 0.51 | 0.44-0.59 | <0.001 | 0.85 | 0.71-1.02 | 0.074 |
| **Writing** |  |  |  |  |  |  |
| Monthly | 0.97 | 0.88-1.08 | 0.577 | 0.89 | 0.79-1.01 | 0.063 |
| Weekly | 1.01 | 0.91-1.13 | 0.857 | 1.00 | 0.88-1.14 | 0.980 |
| **Baking/cooking** |  |  |  |  |  |  |
| Monthly | 0.94 | 0.85-1.03 | 0.193 | 0.90 | 0.80-1.02 | 0.100 |
| Weekly | 0.87 | 0.78-0.96 | 0.004 | 0.85 | 0.75-0.95 | 0.005 |
| **Sewing** |  |  |  |  |  |  |
| Monthly | 1.25 | 1.07-1.46 | 0.005 | 1.02 | 0.85-1.23 | 0.796 |
| Weekly | 1.25 | 1.06-1.47 | 0.007 | 0.89 | 0.74-1.06 | 0.196 |
| **Hobby/project** |  |  |  |  |  |  |
| Monthly | 0.73 | 0.66-0.81 | <0.001 | 1.01 | 0.90-1.14 | 0.877 |
| Weekly | 0.59 | 0.53-0.65 | <0.001 | 0.81 | 0.71-0.92 | 0.001 |
| **Club** |  |  |  |  |  |  |
| Monthly | 0.67 | 0.61-0.74 | <0.001 | 0.82 | 0.73-0.93 | 0.001 |
| Weekly | 0.61 | 0.54-0.69 | <0.001 | 0.82 | 0.71-0.94 | 0.005 |
| **Organization** |  |  |  |  |  |  |
| Monthly | 0.84 | 0.75-0.95 | 0.005 | 0.86 | 0.75-0.99 | 0.041 |
| Weekly | 1.15 | 0.94-1.4 | 0.166 | 1.09 | 0.87-1.38 | 0.442 |

*Note.* N=16,043. For all activities, no engagement was the reference category. Model 2 was adjusted for depression at the previous wave, gender, race/ethnicity, education, age, marital status, employment status, household income, neighborhood safety, social network, difficulty with ADLs and IADLs, chronic health conditions, and cognition. Results weighted and based on 20 multiply imputed data sets. Bold text indicates $p < 0.05$ .

### Sensitivity analyses

After adjusting for confounders, we found no evidence for interactions between any activities and gender on depression (Table S5). Concurrently, there was also no evidence for interactions between any activities and age group on depression (Table S6). Longitudinally, there was initially evidence that the association between writing and depression differed according to age group, but this did not survive adjustment for confounders (Table S6). In the adjusted longitudinal models, there was only evidence for an interaction between baking/cooking something special and age group on subsequent depression. For those aged 50-59 years, baking/cooking something special weekly was associated with 30% (95% CI=0.56-0.87) lower odds of subsequent depression. Baking/cooking something special was not associated with subsequent depression in the older age groups.

Using the alternative threshold for the CES-D and repeating analyses using the CIDI-SF did not substantially alter our findings (Tables S7-S8). Although including the preliminary data from the 2018 HRS wave did not alter most findings, there was additional evidence that reading monthly and weekly and baking/cooking something special weekly were associated with lower odds of concurrent depression (Table S9). Longitudinally, there was evidence that weekly reading was associated with lower odds of subsequent depression, although all other findings remained similar (Table S9). Differences in findings may be a result of missing data on confounders in 2018 or biases in sample selection (see Supplement).

## Discussion

We explored the associations between engagement in a diverse range of leisure activities and concurrent and subsequent depression in a large nationally representative cohort of older adults in the US. Attending a sport, social, or other club weekly or monthly was consistently associated with reduced odds of concurrent and subsequent depression. Spending time on a hobby or project weekly was also associated with lower odds of concurrent and subsequent depression, but for monthly hobby or project engagement this association was only present concurrently, not longitudinally. Although baking or cooking something special was not associated with concurrent depression, weekly baking or cooking something special was associated with reduced odds of depression two years later. These associations were independent of a range of confounders and previous depression. However, writing (e.g., letters, stories, or journal entries), reading books magazines or newspapers, making clothes knitting or embroidery (sewing), and attending non-religious organizations (political, community, or other interest groups) were not consistently associated with depression. Additionally, there was no evidence that the association between hobbies or projects, sport social or other clubs, and depression varied according to gender or age group. However, more frequent baking or cooking something special was only associated with lower subsequent depression in those aged 50-59.

Both overall engagement in participatory leisure activities and attending cultural events have previously been shown to be associated with a reduced risk of subsequent depression in older adults (Fancourt and Steptoe, 2019; Fancourt and Tymoszuk, 2019; Glass et al., 2006; Katz and Yelin, 2001; Lampinen et al., 2006). We aimed to build on this evidence by exploring whether specific participatory activities, with diverse characteristics, were differentially related to depression. We were particularly interested in whether the extent to which activities involved creative or artistic elements might alter their influence on depression in older adults. Of the activities we included, those most often referred to as the arts (writing letters stories or journal entries, reading books magazines or newspapers, sewing [making clothes knitting or embroidery]) were not associated with depression. We therefore found no evidence that more artistic or creative activities were associated with depression. This is in contrast to previous evidence that creative activities provide additional benefits for older adults over other leisure activities (Dadswell et al., 2017; Fancourt et al., 2021) and the positive effects of creative arts interventions for older adults experiencing depression (Chen et al., 2021; Curtis et al., 2018; Dunphy et al., 2019; Fancourt et al., 2020a, 2020b; Zhao et al., 2016). However, it has been proposed that the concept of creativity should not be limited to formal arts activities and should instead be extended to being creative in daily life, including in constructing personal knowledge and understanding, problem solving, and performing everyday tasks in an imaginative or artistic manner (Beghetto and Kaufman, 2007; Bellass et al., 2019). Using this definition, one could argue that all leisure activities included in this study required creativity, and the extent to which activities have traditionally been included in definitions of the arts becomes less relevant.

The leisure activities included in this study also differed in their specificity. Four of the activities were well-defined, included a relatively narrow range of behaviors, and were likely to take place in the home. These were reading (books, magazines, or newspapers), writing (e.g., letters, stories, or journal entries), baking or cooking something special, and sewing (making clothes, knitting, embroidery). In contrast, the other three activities (doing hobbies or projects, sport social or other clubs, and non-religious organizations) were quite broad and could have included a range of behaviors in a variety of settings. Of these activities, three of the four specific items were not associated with depression, whereas most of the more general leisure activities were. These findings are consistent with existing evidence that engagement in broadly defined hobbies is associated with lower odds of depression (Dupuis and Smale, 1995; Fancourt et al., 2020a; Hirosaki et al., 2009). This makes it difficult to study the specific mechanisms underlying the association between participation in leisure activities and depression and suggests that the mechanisms are likely to differ across individuals (Fancourt et al., 2021). Future research could further investigate whether it is the way in which leisure activities are defined that affect associations with depression, the settings in which activities occur, or other factors that are most important.

It is widely accepted that leisure activities that involve social interactions are particularly beneficial for mental health in older age (Adams et al., 2011; Dadswell et al., 2017; Dupuis and Alzheimer, 2008; Ryu and Heo, 2018). In line with this, we found that attending sport, social, or other clubs was most strongly associated with depression. However, attending non-religious organizations (political, community, or other interest groups) was not consistently associated with lower odds of depression. These contrasting relationships demonstrate the importance of investigating leisure activities separately. To understand these differences, we can consider the potential active ingredients of attending sport, social, or other clubs, which may activate causal mechanisms that reduce depression (Fancourt et al., 2021). For example, one active ingredient of clubs is the provision of social resources that can reduce loneliness and can be used in the development of a social identity (Dadswell et al., 2017; Haslam et al., 2009). Social identity theory states that individuals build a sense of who they are based on their group membership (Hogg, 2006), and there is evidence that individuals with stronger social identities have lower rates of depression (Cruwys et al., 2014; Haslam et al., 2009; Postmes et al., 2019). Although one could expect similar active ingredients of non-religious organizations (political, community, or other interest groups), it is possible that these more formal groups might not lead to the formation of strong social identities or positive social interactions, and thus are not associated with depression.

Although attending a sport, social, or other club was most consistently associated with depression, there were still associations between doing hobbies or projects, baking or cooking something special, and depression after accounting for the association between clubs and depression, suggesting that these activities provide additional active ingredients. They are not necessarily social activities, although they could lead to social interactions. By providing cognitive stimulation, hobbies or projects and baking or cooking something special could activate cognitive mechanisms including enhanced cognitive restructuring (identifying problematic cognitions and rationalizing and rebutting these thoughts; Fancourt et al., 2021). This is a key element of treatments for depression, such as cognitive behavioral therapy (CBT), and is effective in reducing depressive symptoms (Clark, 2013). As more sedentary leisure activities (writing, reading, sewing) were not associated with depression, another important mechanism could be the physical activity involved in the other leisure activities (Dunphy et al., 2019; Fancourt et al., 2021). Increased physical activity and fitness is protective against depression in older adults (Kandola et al., 2020, 2019; Schuch et al., 2018). There may also be other active ingredients of clubs, hobbies or projects, and baking or cooking something special, leading to further mechanisms that reduce depression, which may interact and operate cyclically (Fancourt et al., 2021). These mechanisms should be explored further in longitudinal population-based studies.

In contrast to previous evidence for gender-dependent associations in working age adults (Cuypers et al., 2012), we found no evidence that engagement in leisure activities differentially affected depression in men and women. Activities may be equally beneficial across genders, even if there are gender differences in engagement. We did not expect to find age differences in the association between baking or cooking something special and depression. It is possible that adults aged 50-59 were more likely to bake or cook for friends or family, and thus more frequent engagement in this activity was an indicator of increased social interactions in this age group, which were then associated with reduced odds of subsequent depression (Kawachi and Berkman, 2001). However, adjusting for social network composition did not attenuate this association.

### Strengths and limitations

This study has several strengths. HRS is a large nationally representative cohort of older adults that allowed us to investigate population-averaged concurrent and longitudinal associations between leisure activities and depression, whilst controlling for previous depression and a range of confounders. Using alternative indicators of depression in sensitivity analyses did not alter our findings. However, the way in which questions were asked in HRS limits investigation of demographic factors and makes the extent of creativity involved in leisure activities unclear. Sport, social, or other clubs and hobbies or projects may or may not have involved creative activities. The social nature of activities was not recorded, so we could not explore this further. Additionally, there are a wide range of cultural, artistic, and creative leisure activities that were not measured in HRS, including activities such as music making and listening, dance, and visual arts. Future research should investigate the associations between participating in these activities and depression in more detail. Although we adjusted for a range of confounders, residual confounding is possible, as there is a social gradient in engagement in leisure activities and depression (Bone et al., 2021; Fluharty et al., 2021). It remains difficult to disentangle whether the association between engagement and depression is due to self-selection, reverse causality, or because these activities reduce depression. We also were not able to adjust for average engagement in leisure activities at the neighbourhood level (as these data were not available in HRS), which could have influenced the associations between leisure activity engagement and depression (Adjaye-Gbewonyo et al., 2018). Additionally, we recognize that gender is not a binary construct, although we had to treat it as such. Further, as HRS combined a range of races/ethnicities into the Other race/ethnicity category, we were not able to investigate the influence of race/ethnic identities, and associated racism and cultural caste systems. By exploring the population-level associations between leisure activities and depression, we conflated individual experiences, so future research should focus on the needs of different groups, who may experience distinctive stressors and outcomes (Fernando, 1984; Williams, 2018).

## Conclusions

Our findings indicate that engagement in some leisure activities, which may be more accessible to older adults than receptive cultural engagement, are independently associated with reduced odds of depression. However, we found no evidence that more artistic or creative activities were associated with depression. Despite this, and given the high prevalence of depression in older adults, the fact that is commonly undiagnosed and untreated, the impacts on daily life, and comorbidity with other health problems (Rodda et al., 2011; World Health Organization, 2015), it is promising that engagement in sport social or other clubs, hobbies or projects, and baking or cooking something special could lead to lower rates of depression. These associations may occur through the activation of mechanisms such as increased social identity, cognitive stimulation, and physical activity. Associations persist across genders and age groups, holding promise for many older adults in the US. The potential of these easily accessible activities to reduce depression is particularly important in the wake of COVID-19 with the closure of many arts venues. It is possible that an individual’s own choices of activities, be it sport social or other clubs, hobbies or projects, or baking or cooking something special, are more beneficial for their mental health than creative arts interventions that often only provide the opportunity to engage in a specific activity (Curtis et al., 2018; Dunphy et al., 2019). Policy makers should consider how older adults can be supported to engage in leisure activities that may help to prevent depression as they age. For example, in the United Kingdom, social prescribing systems allow primary care practitioners to connect older adults to community groups, which may improve health (Drinkwater et al., 2019). Similar systems could be tested to establish whether they have the potential to reduce the risk of depression in older adults in the US.

## Data Availability

In this study, we combined several HRS public use datasets (RAND HRS Longitudinal File 2016 (V2); RAND HRS Detailed Imputations File 2016 (V2); RAND HRS 2008-2016 Fat Files). This data is all freely available from the HRS website.

https://hrs.isr.umich.edu/data-products

## Acknowledgements

We thank Shanae Burch, thought leader on work at the intersections of the arts, equity, and public health in the US, for her comments on this manuscript. We also gratefully acknowledge the contribution of the HRS study participants.

## Notes

Funding The EpiArts Lab, a National Endowment for the Arts Research Lab at the University of Florida, is supported in part by an award from the National Endowment for the Arts (1862896-38-C-20). The opinions expressed are those of the authors and do not represent the views of the National Endowment for the Arts Office of Research & Analysis or the National Endowment for the Arts. The National Endowment for the Arts does not guarantee the accuracy or completeness of the information included in this material and is not responsible for any consequences of its use. The EpiArts Lab is also supported by the University of Florida, the Pabst Steinmetz Foundation, and Bloomberg Philanthropies. This work was also supported by an award from Arts Council England (INVF-00404365). DF is supported by the Wellcome Trust (205407/Z/16/Z).

Declaration of interest No authors report any conflicts of interest.

### Competing Interest Statement

The authors have declared no competing interest.

### Funding Statement

The EpiArts Lab, a National Endowment for the Arts Research Lab at the University of Florida, is supported in part by an award from the National Endowment for the Arts (1862896-38-C-20). The opinions expressed are those of the authors and do not represent the views of the National Endowment for the Arts Office of Research & Analysis or the National Endowment for the Arts. The National Endowment for the Arts does not guarantee the accuracy or completeness of the information included in this material and is not responsible for any consequences of its use. The EpiArts Lab is also supported by the University of Florida, the Pabst Steinmetz Foundation, and Bloomberg Philanthropies. This work was also supported by an award from Arts Council England (INVF-00404365). DF is supported by the Wellcome Trust (205407/Z/16/Z).

### Author Declarations

This study has Institutional Review Board approval from the University of Florida (IRB201901792) and ethical approval from University College London Research Ethics Committee (project 18839/001).

### Summary of Updates

Manuscript updated following peer review

